# The Pattern of rpoB gene mutation of *Mycobacterium tuberculosis* and predictors of rifampicin resistance detected by gene Xpert MTB/Rif in Tanzania

**DOI:** 10.1101/2023.12.23.23300496

**Authors:** Peter Richard Torokaa, Heledy Kileo, Loveness Urio, Mariam R. Mbwana, Mariam C. Monah, Sephord Saul Ntibabara, Jasper Kimambo, Paschal Seleman, Collins Franklin, Robert Balama, Riziki M. Kisonga, Mtebe V. Majigo, Agricola Joachim

**Affiliations:** Muhimbili University of Health and Allied Sciences, School of Public Health and Social Sciences, Dar es Salaam, Tanzania; Tanzania Field Epidemiology and Laboratory Training Program, Dar es Salaam, Tanzania; Ministry of Health, National TB and Leprosy Program, Dodoma Tanzania; Muhimbili University of Health and Allied Sciences, College of Medicine, Dar es Salaam, Tanzania

**Author notes:** Corresponding Author: Peter Torokaa, (PRT).

**Keywords:** *Mycobacterium tuberculosis*, rifampicin resistance, GeneXpert, probe mutations, Tanzania

## Abstract

**Introduction:** Antimicrobial resistance associated with *Mycobacterium tuberculosis (*MTB) is the challenge facing Tuberculosis (TB) management worldwide. Rifampicin resistance (RR) has been associated with the rpoB gene mutation. No study was conducted in Tanzania to determine the commonest mutation. The inconsistent findings from various studies support the need to determine whether reported mutation patterns are applicable in our setting. We determined the frequency of rpoB gene mutation and factors associated with RR detected using GeneXpert MTB/Rif.

**Methods:** We conducted a retrospective cross-sectional study involving data from the National Tuberculosis and Leprosy Program database from 2020 to 2022 for cases investigated using GeneXpert. Descriptive analysis was performed as the frequency for categorical variables. The chi-square test and regression analysis assessed the relationship between the independent variables and outcome. The 95% confidence interval and a significance level of p<0.05 were used to assess the association strength.

**Results:** A total of 56,004 participants had status of MTB and RR. Most, 38,705/56,004 (69.11%) were male. Probe E, 89/219 (40.64%) was the predominant. Human immunodeficiency virus (HIV)-positive patients had a higher gene mutation, 134/10601 (1.26%) than HIV-negative, 306/45016 (0.68%) (p<0.001). Patients with both pulmonary and extra-pulmonary TB had about four times greater odds of developing rifampicin resistance (AOR 3.88, 95%CI: 1.80 – 8.32). RR was nearly nine times higher in previously treated patients than new patients (AOR 8.66, 95%CI: 6.97–10.76). HIV-positive individuals had nearly twice the odds of developing RR than HIV-negative individuals (AOR 1.91, 95%CI: 1.51 – 2.42).

**Conclusion:** The rate of RR was low compared to other studies in Tanzania, with probe E mutations the most prevalent. Patients with disseminated TB, HIV co-infection and those with prior exposure to anti-TB had more risk of RR. The findings highlight the need to strengthen surveillance of multidrug-resistant-TB among patients with a higher risk of RR.

## Introduction

Multidrug-resistant *Mycobacterium tuberculosis* (MDR-TB) is the biggest challenge facing Tuberculosis (TB) care and treatment worldwide (1). MDR-TB is a major contributor to global antimicrobial resistance (AMR) and continues to pose a public health threat (1–3). MDR-TB is considered when there is resistance to isoniazid and rifampicin, with or without resistance to other first-line drugs. The main problem is rifampicin resistance, the most potent first-line treatment (1). Patients with rifampicin-resistant TB or MDR-TB require a second-line treatment regimen (4). Globally, the annual estimated number of MDR-TB or rifampicin-resistant TB cases was steady between 2015 and 2020; however, it increased in 2021. In 2021, it was estimated to be 450,000 events, a 3.1% increase from 437,000 in 2020 (1).

The World Health Organization (WHO) developed a strategy to end TB-related deaths, illnesses, and suffering by 2035 (5). It recommends accelerating the identification and enhancing treatment for MDR-TB. Apart from access to diagnosis, adequate infection control must also be implemented in the settings where patients are treated (4). To diagnose MDR-TB, WHO recommends more specific and sensitive assays like real-time polymerase chain reaction (RT-PCR), DNA microarray, and loop-mediated isothermal amplification (LAMP). These technologies determine the mutations in the genes to identify drug resistance. In some populations of HIV-infected presumptive TB patients, the lateral flow lipoarabinomannan assay (LF-LAM) test is advised to aid in the diagnosis of TB (6).

The invention of the Xpert® MTB/RIF Assay (Cepheid, USA), which simultaneously detects the presence of *Mycobacterium tuberculosis* (MTB) and rifampicin resistance, has transformed the diagnosis of TB. Rifampicin inhibits DNA-directed RNA synthesis of MTB proteins by binding to the subunit of the bacterial DNA-dependent RNA polymerase *(rpoB*) enzyme. Rifampicin resistance has been associated with the rpoB gene mutation. Almost 96.1% of rifampicin-resistant MTB strains have *rpoB* mutations (7). The rifampicin resistance is the proxy sign of the MDR-TB (8,9). GeneXpert/MTB RIF is a cartridge-based, automated hemi-nested-nested real-time Polymerase Chain Reaction (PCR) system that utilizes five overlapping probes named Probe A (codons 507–511), Probe B (codons 511–518), Probe C (codons 518–523), Probe D (codons 523– 529) and Probe E (codons 529–533) (10).

Studies in Africa revealed that the commonest mutation of rpoB occurs at probes E and D (11–13). However, the literature shows different pattern distributions of the rpoB gene mutation of MTB associated with drug resistance (11–13). There is a need to determine whether the findings in other studies apply to our setting due to the varied findings obtained in various geographical areas. No other study has been conducted in Tanzania to reveal the patterns of rpoB mutation. Due to efforts to increase access to GeneXpert MTB/RIF diagnosis in the country, there is also a need to determine the factors associated with Rifampicin-resistant TB.

## Methods

### Study design and setting

We conducted a retrospective cross-sectional study involving the Tanzania National Tuberculosis and Leprosy Program (NTLP) data collected from January 2020 through December 2022. The study covered Presumptive TB cases notified to NTLP from 26 regions of Tanzania’s mainland. The NTLP is tasked with preventing tuberculosis and leprosy as serious public health concerns in Tanzania, and it was launched in 1977 as a single combined programme for the two diseases.

### Data collection

We extracted demographic and clinical data from eTL and laboratory results from gxalert databases on 10^th^ October, 2023 and then exported them to Microsoft Excel. The demographic data from the eTL register and laboratory results data were linked between these two databases. The identifiers used to link the two databases were patient name, health facilities, district, and year. The variables of interest from the two databases were demographic characteristics (age, gender, residence), clinical characteristics (TB treatment history, type of TB, bacillary load, HIV status), and geneXpert results.

### Data analysis

We presented the descriptive analysis with frequency distributions (%) for categorical variables and a mean was a measure of central tendency for age. Regression analyses (bivariate and multivariate) were performed to assess the relationship between demographic and clinical characteristics as independent variables and the rifampicin resistance as an outcome variable. Factors with a p-value of 0.20 in the bivariate analyses were included in the multivariable model using forward selection. The 95% confidence interval (CI) was presented, and a significance level of p <0.05 was used.

## Results

We extracted data for 112,768 nonrepeating patients tested for TB using GeneXpert. Of all, 56,662 (50.2%) had laboratory-confirmed TB. Rifampicin resistance was identified in 450/56,662 (0.8%) patients with MTB. The specific gene mutations in the rpoB gene of *Mycobacterium tuberculosis* were identified in the form of the missing probes, which indicated a specific mutation occurs at a specific codon in the rpoB gene named Probe A (codons 507–511), Probe B (codons 511–518), Probe C (codons 518–523), Probe D (codons 523–529), and Probe E (codons 529–533). Missing probes (specific rpoB mutation detected) were found in 219/ 450 (48.7%) samples with Rifampicin-resistant MTB, and no missing probes (no specific rpoB mutation detected) in 231/450 (51.3%) with Rifampicin-resistant. Rif Indeterminate were observed in 658/56,662 (1.16%) of patients with MTB and were excluded in further analysis (Fig 1).

**Fig 1.**
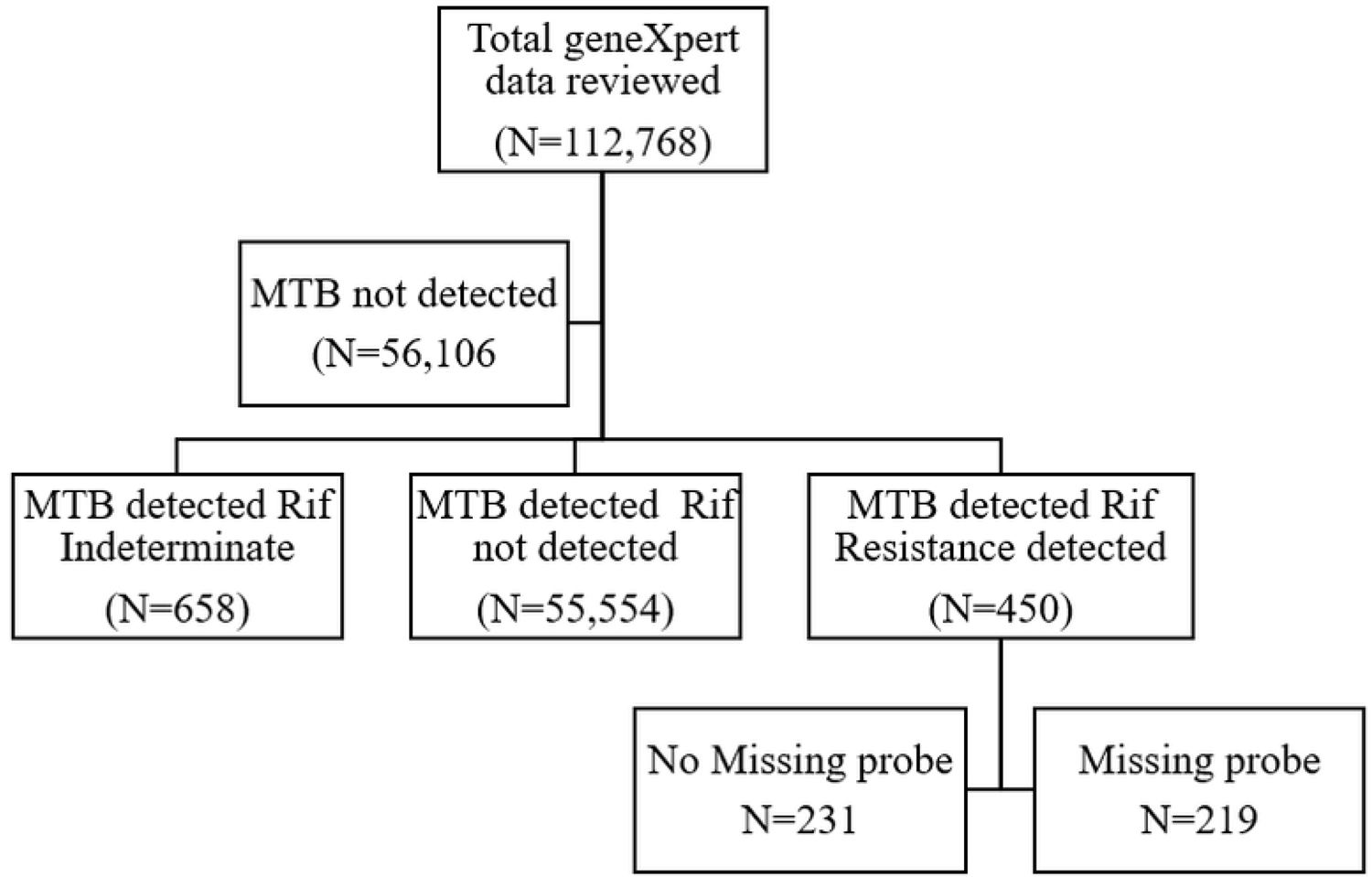
Flow chart of participants’ analysis.

### Demographic and clinical characteristics of study participants

A total of 56,004 participants had clear status of MTB and rifampicin resistance. Most of them, 38,705/56,004 (69.11%) were male. The age group 35-44 years accounted for the largest cases, 13,512/56,004 (24.13%). Most participants, 55,192/56,004 (98.55%), had pulmonary TB. Participants with new TB were 53,818/56,004 (96.1%), and 45,016/56,004 (80.38%) were HIV-negative (Table 1). The Dar es Salaam region contributed 10,524/56,004 (18.79%) cases, which is higher than in any other region. The lowest number of cases were reported from the Katavi region. The regional distribution of cases is shown in Fig 2.

**Fig 2.**
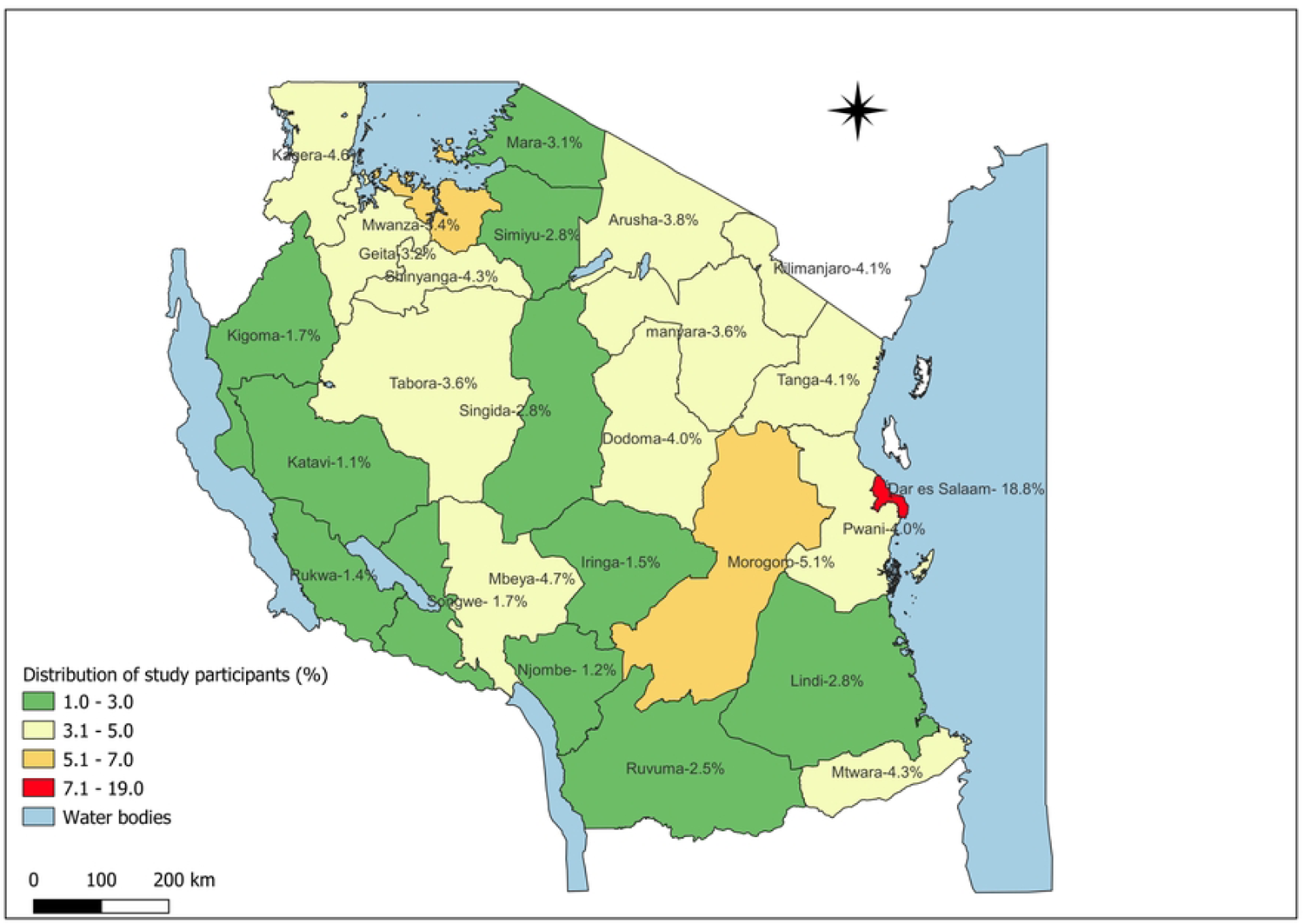
Map showing the proportion distribution of study participants per region.

**Table 1.** Participants’ demographic and clinical characteristics (N=56,004)

| Variable | Frequency | Percent (%) |
| --- | --- | --- |
| **Age Group (years)** |  |  |
| <15 | 1765 | 3.15 |
| 15-24 | 6441 | 11.5 |
| 25-34 | 12637 | 22.56 |
| 35-44 | 13512 | 24.13 |
| 45-54 | 10460 | 18.68 |
| >55 | 11189 | 19.98 |
| Mean age (SD) | 41(16.4) |  |
| **Gender** |  |  |
| Male | 38705 | 69.11 |
| Female | 17299 | 30.89 |
| **TB Type** |  |  |
| Pulmonary | 55192 | 98.55 |
| Extra pulmonary | 550 | 0.98 |
| Pulmonary and extra pulmonary | 262 | 0.47 |
| **Treatment History** |  |  |
| New patient | 53818 | 96.1 |
| Previous treated | 2186 | 3.9 |
| **HIV status** |  |  |
| Positive | 10601 | 18.93 |
| Negative | 45016 | 80.38 |
| Unknown | 387 | 0.69 |

### Pattern of specific rpoB gene mutations in MTB

Specific mutations (missing probe) were detected in 219/450 (48.7%) rifampicin-resistant TB. The most prevalent mutation, 89/219 (40.64%), occurred at Probe E, followed by Probe D, 44/219 (20.09%), and the least mutation, 10/219 (4.56%), occurred at Probe C. We found a mutation combination of Probe A and B, 5/219 (2.28%), Probe A and D, 2/219 (0.91%), Probe A and E 1/219 (0.46%), and Probe B and E 1/219 (0.46%). One patient had a triple mutation combination at Probe A, D, and E 1/219 (0.46%) (Table 2).

**Table 2.** Frequency and distribution of specific rpoB gene mutations (N=219)

| Probes | #Codons | Frequency | Percent (%) |
| --- | --- | --- | --- |
| Probe A | 507–511 | 21 | 9.59 |
| Probe B | 511–518 | 39 | 17.81 |
| Probe C | 518–523 | 10 | 4.57 |
| Probe D | 523–529 | 44 | 20.09 |
| Probe E | 529–533 | 89 | 40.64 |
| Probe A+B |  | 5 | 2.28 |
| Probe A+D |  | 2 | 0.91 |
| Probe A+E |  | 1 | 0.46 |
| Probe B+E |  | 1 | 0.46 |
| Probe A+D+E |  | 1 | 0.46 |

### The proportion of participants with no missing probe

Specific rpoB mutation were not detected (no missing probe) in 231/450 (51.3%) participants with Rifampicin-resistance. We found that no missing probes were significantly higher in new TB patients, 189/334 (56.59%) compared to previously treated patients, 42/116(36.21%) (p = 0.003). Patients with a very low TB bacillary load, 199/263 (75.67%), had a higher number of no missing probes when compared with other categories (p<0.001). There was no statistically significant difference of no missing probes for other variables (Table 3).

**Table 3.**
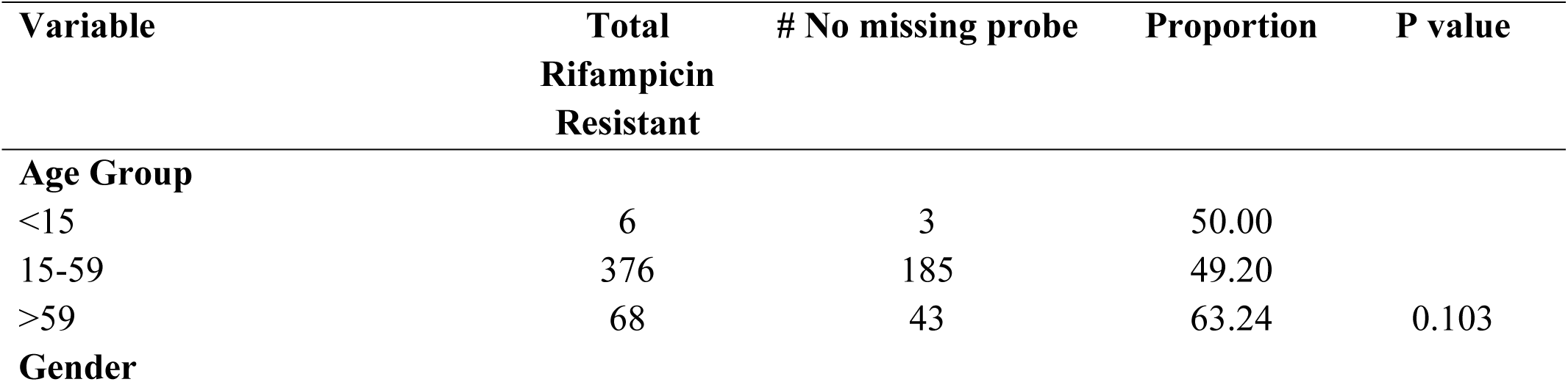
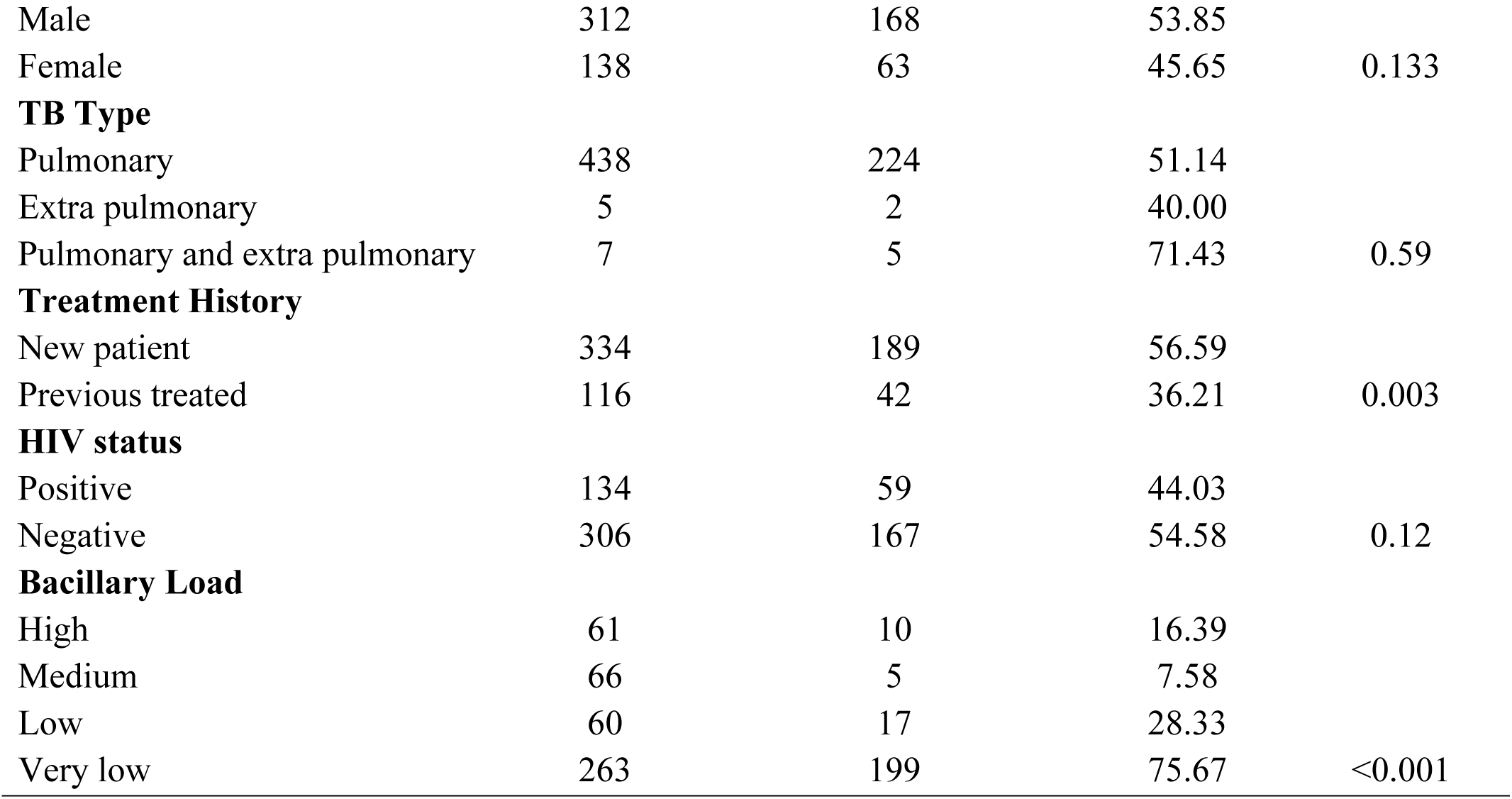
Demographic and Clinical characteristics of the number of no-missing probe study participants (N=450)

### Rifampicin resistance and participant characteristics

GeneXpert detects the mutation in the rpoB mutations, and the final results are provided as rifampicin-resistant. The rpoB gene mutation was higher in 15-59-year-old patients, 383/45,782 (0.84%) compared to other age groups (p = 0.047). More mutations were found in patients with pulmonary and extra-pulmonary TB patients (7/262, 2.67%) than in pulmonary or extra-pulmonary alone (p = 0.003). Furthermore, we found that previously treated patients had a higher mutation rate, 116/2186 (5.31%), compared to new patients (p<0.001). HIV-positive TB patients had a higher gene mutation, 134/10601 (1.26%), than HIV-negative, 306/45016(0.68%) (p<0.001). Other variables like gender and point of health service showed no significant difference in gene mutation (Table 4).

**Table 4.** Distribution of rifampicin resistance and participants’ characteristics (N=56,004))

| Variable | Total participants | # Total RR | Proportion | P value |
| --- | --- | --- | --- | --- |
| **Age Group** |  |  |  |  |
| <15 | 1765 | 6 | 0.34 |  |
| 15-59 | 45782 | 383 | 0.84 |  |
| >59 | 8457 | 61 | 0.72 | 0.047 |
| **Gender** |  |  |  |  |
| Male | 38705 | 312 | 0.81 |  |
| Female | 17299 | 138 | 0.80 | 0.918 |
| **TB Type** |  |  |  |  |
| Pulmonary | 55192 | 438 | 0.79 |  |
| Extra pulmonary | 550 | 5 | 0.91 |  |
| Pulmonary and extra pulmonary | 262 | 7 | 2.67 | 0.003 |
| **Treatment History** |  |  |  |  |
| New patient | 53818 | 334 | 0.62 |  |
| Previous treated | 2186 | 116 | 5.31 | <0.001 |
| **HIV status** |  |  |  |  |
| Positive | 10601 | 134 | 1.26 |  |
| Negative | 45016 | 306 | 0.68 | <0.001 |
| **Referral type** |  |  |  |  |
| Community | 46897 | 364 | 0.78 |  |
| Health facility | 6167 | 60 | 0.97 |  |
| Others | 2940 | 26 | 0.88 | 0.234 |

\*\* The frequency for HIV status is less due to unknown status.

### Predictors of rifampicin resistance

Compared to pulmonary TB, patients with both pulmonary and extra-pulmonary TB had about four times greater odds of developing rifampicin resistance (AOR 3.88, 95%CI: 1.80 – 8.32). The likelihood of rifampicin resistance was nearly nine times higher in previously treated patients compared to new (AOR 8.66, 95%CI: 6.97 – 10.76). HIV-positive individuals had nearly twice the odds of developing rifampicin resistance than HIV-negative individuals (AOR 1.91, 95%CI: 1.51 – 2.42) (Table 5).

**Table 5.** Multivariate Logistic regression analysis for the predictors of Rifampicin resistance.

| Characteristic | Rif. resistance<br>n (%) | COR (95% CI) | P-value | AOR (95% CI) | P-value |
| --- | --- | --- | --- | --- | --- |
| **Age Group** |  |  |  |  |  |
| <15 | 6 (0.34) | Ref |  | Ref |  |
| 15-59 | 383 (0.84) | 2.47 (1.10 – 5.55) | 0.028 | 2.07 (0.92 – 4.67) | 0.078 |
| >59 | 61 (0.72) | 2.13 (0.92 – 4.93) | 0.078 | 1.95 (0.84 – 4.54) | 0.119 |
| **TB Type** |  |  |  |  |  |
| Pulmonary | 438 (0.79) | Ref |  | Ref |  |
| Extra pulmonary | 5 (0.91) | 1.15 (0.47 – 2.78) | 0.762 | 1.19 (0.49 – 2.92) | 0.694 |
| Pulmonary and extra pulmonary | 7 (2.67) | 3.43 (1.61 -7.31) | 0.001 | **3.88 (1.80 – 8.32)** | **0.001** |
| **Treatment History** |  |  |  |  |  |
| New patient | 334 (0.62) | Ref |  | Ref |  |
| Previous treated | 116 (5.31) | 8.97 (7.23 – 11.13) | <0.001 | **8.66 (6.97 – 10.76)** | **<0.001** |
| **HIV status** |  |  |  |  |  |
| Negative | 306 (1.26) | Ref |  | Ref |  |
| Positive | 134 (0.68) | 1.87 (1.53 – 2.29) | <0.001 | **1.82 (1.44 – 2.30)** | **<0.001** |
| **Referral type** |  |  |  |  |  |
| Community | 364 (0.78) | Ref |  | Ref |  |
| Health facility | 60 (0.97) | 1.25 (0.95 – 1.65) | 0.104 | 0.83 (0.61 – 1.14) | 0.250 |
| Others | 26 (0.88) | 1.14 (0.76 – 1.70) | 0.519 | 0.99 (0.66 – 1.48) | 0.969 |

## Discussion

The current study found that the most prevalent mutation in rpoB gene mutation in MTB was at probe E codon (529–533) (40.64%), followed by Probe D codon (523–529) 20.09% which was similar to studies done in Nigeria, Uganda, Pakistan, Addis Ababa Ethiopia, and Bangladesh (13–17), the similarities suggests that a significant number of low-income individuals migrating for shelters may be responsible for the spread of mutant strains within the community (18). Our findings differed from the study done in Northeast India and Enugu, South Eastern Nigeria, which revealed that the most prevalent mutations were detected at probes A and D, respectively (19,20). The differences might be attributed to geographical differences caused by the variations in the *M. tuberculosis* lineage (21). According to the results of our study and previous studies, probe E is linked to the most common probe mutation in the rpoB gene mutation.

The study found that new TB patients had a significantly higher number of specific rpoB mutations not detected among those with rifampicin resistance than previously treated patients. Our finding is similar to a Northeast India study (20). This may result from an early stage of TB diagnosis, which may have a low bacillary load. No missing probe may also be triggered by various probes having varying target hybridization dynamics, which might have a higher impact after extended PCR cycles (22). Patients with low TB bacillary load also had more no-missing probes. This could mean GeneXpert testing in patients with less advanced disease and lower bacterial density (new patients) is considered less accurate in detecting rifampicin-resistant tuberculosis (23). The very low bacillary load on GeneXpert testing was shown to be substantially linked to false rifampicin resistance during the initial GeneXpert assay (22). The samples with very low bacillary load were not retested; hence, the rate of false positives for rifampicin resistance could not be determined.

The rifampicin resistance was found more in patients with both pulmonary and extra-pulmonary TB than in pulmonary or extra-pulmonary alone. In addition, patients with both pulmonary and extra-pulmonary TB had almost four times greater odds of developing rifampicin resistance. Our findings were comparable to the study done in the Debre Markos Referral Hospital Ethiopia (24). We also found that previously treated patients had significantly higher rifampicin resistance than new patients. The likelihood of rifampicin resistance was nearly nine times higher in previously treated patients than in new patients, similar to the study done in Nepal (25). Our findings support that MTB increases the ability to develop resistance when exposed to anti-TB drugs, especially in patients with poor adherence to treatment (26). In our study, the odds of developing rifampicin resistance in previously treated patients were higher than in Somalia and other East Gojjam zone northwest Ethiopia studies, which revealed four and six times, respectively (27,28).

HIV-positive TB patients had a higher gene mutation than HIV-negative. HIV-positive individuals had nearly twice the odds of developing rifampicin resistance than HIV-negative individuals; this was similar to the studies done in northwest Ethiopia (27,29). Variations in TB/HIV co-infection have been reported in several studies, indicating challenges in diagnosis and treatment due to unusual clinical presentations and difficulties in diagnosis and treatment (30–32). This may be due to differences in TB control strategies and approaches (29). More rifampicin-resistant MTB in HIV patients might be caused by poor treatment adherence. Poor adherence to treatment is the primary factor contributing to drug resistance (25,31). This highlights the need for more surveillance and community involvement.

The study had limitations, including missing variables and being unable to confirm false positive rates for rifampicin resistance at low bacillary load. However, the large sample size of participants from all Tanzania mainland regions made the findings representative and generalisable to all regions.

## Conclusion

The rate of rifampicin resistance in our study was low compared to other studies done in Tanzania. The Probe E (codons 529–533)-related mutations were the most prevalent rpoB gene mutation. Patients with disseminated TB, HIV co-infection and those with prior exposure to anti-TB are associated with rifampicin resistance. The findings highlight the need to strengthen the surveillance of MDR-TB among patients identified with a higher risk of rifampicin resistance.

## Data Availability

All data produced in the present study are available upon reasonable request to the authors

## Ethical considerations

Ethical approval was obtained from the MUHAS Senate Research and Publications Committee of Muhimbili University of Health and Allied Sciences, Ref. No. DA.282/298/01L/629. Approval to use the national TB program data was obtained from the NTLP Program Manager in the Ministry of Health (MoH). The data was routinely collected by health facilities providing TB care and treatment services in which ethical issues are strongly advocated.

## Author contributions

PRT, AJ, HK, MVM and LU authors contributed equally to the design. PRT, MRM, MCM, SSN, RMK, CF, RB, PS and JK did data collection and write-up of this manuscript. AJ and MVM reviewed the manuscript. All authors read and approved the final manuscript.

## Acknowledgments

We are very grateful to the Muhimbili University of Health and Allied Sciences, Ministry of Health, NTLP program, Tanzania Field Epidemiology and Laboratory program for technical support of this study.

